# USING PHYLOGENETICS TO INFER HIV-1 TRANSMISSION DIRECTION BETWEEN KNOWN TRANSMISSION PAIRS

**DOI:** 10.1101/2021.05.12.21256968

**Authors:** Christian Julian Villabona-Arenas, Stéphane Hué, James A. C. Baxter, Matthew Hall, Katrina A. Lythgoe, John Bradley, Katherine E. Atkins

## Abstract

Inferring the transmission direction between linked individuals living with HIV provides unparalleled power to understand the epidemiology that determines transmission. Phylogenetic ancestral state reconstruction approaches infer the transmission direction by identifying the individual in whom the most recent common ancestor of the virus populations originated. However, these methods vary in their accuracy but it is unclear why. To evaluate the performance of phylogenetic ancestral state reconstruction, we inferred the transmission direction for 112 HIV transmission pairs where transmission direction was known and detailed additional information was available. We then fit a statistical model to evaluate the extent to which epidemiological, sampling, genetic and phylogenetic factors influenced the outcome of the inference. We repeated the analysis under real-life conditions with only routinely-collected data. We found that the inference of transmission direction depends principally on the topology class and branch length characteristics of the phylogeny. Under real-life conditions, the probability of identifying the correct transmission direction increases from 52%—when a monophyletic-monophyletic or paraphyletic-polyphyletic tree topology is observed, when the sample size in both partners is small and when the tip closest to the root does not agree with the state at the root—to 93% when a paraphyletic-monophyletic topology is observed, when the sample size is large and when the tip closest to the root agrees with root state. Our results suggest that discordance between previous studies in inferring the transmission direction can be explained by differences in key phylogenetic properties that arise due to different evolutionary, epidemiological and sampling processes.

**Significance Statement:** Identifying the direction of infectious disease transmission between individuals provides unparalleled power to understand infectious disease epidemiology. With epidemiological and clinical information typically unable to distinguish the direction, phylogenetic analysis of pathogen sequence data is an alternative approach. However, when these phylogenetic methods have been implemented, their accuracy is highly variable, and the reasons for this discordance is unknown. Here we analyse sequence data from over 100 pairs of individuals for whom both the direction of transmission of HIV is known and detailed epidemiological and sampling information is available. We find that easily quantifiable phylogenetic characteristics discriminate whether a phylogenetically-inferred transmission direction is correct. Our analysis highlights that phylogenetic approaches are unsuitable for individual-level analysis such as forensic investigations.

## Introduction

Identifying transmission chains via contact tracing is a cornerstone of infectious disease control. It provides an opportunity to test potential cases, treat infections early and break ongoing transmission. Moreover, identifying the transmission direction provides paramount knowledge for understanding risk factors of transmission and susceptibility (1–3), household transmission (4), spread (5–7) and early pathogenesis events (8). Yet inferring transmission direction is challenging. In rare instances, the transmission direction can be inferred from the comparison of symptoms onset time or testing histories of the partners. However, this method is restricted to cases with known contact histories, and for whom other sources of infection can be ruled out, such as for sexually transmitted infections occurring between self-reported sexual partners.

Comparing the ancestral relationship between pathogen genomes sampled from a putative transmission pair has been proposed as a method to identify the transmission direction (9, 10). Specifically, current approaches propose that the transmission direction may be inferred from phylogenies of pathogen sequences sampled from a pair of linked individuals using parsimony-based algorithms (9, 11). Under this framework, the transmitting individual corresponds to the state at the root—i.e., individual A or B—after minimizing the number of state changes along the phylogeny necessary to explain the observed state distribution at the tips. For example, when paraphyly is observed—i.e., when all sequences from one partner form a monophyletic cluster embedded within the pathogen population of the other partner—it would be concluded that the monophyletic clade represents the recipient’s viral population (9). While simulations suggest that using the topology of a phylogeny reconstructed from multiple viral sequences sampled from a known transmission pair can correctly identify the transmission direction, empirical tests of this hypothesis varied in accuracy. For example, one study reported a correct inference of the direction of HIV transmission in 31/32 transmission pairs, with no direction incorrectly inferred (12) but other studies incorrectly identified the direction of HIV transmission in 4/31 couples (13) or 4/36 couples (14). Moreover, how different factors contribute to the success of transmission direction inference remains elusive and thus the likely success of identifying transmission direction in a particular study is not always obvious.

In this study, we analyzed 112 HIV transmission pairs with detailed additional information available, for whom the transmission direction is known and multiple virus sequences are available, to test whether the transmission direction is accurately inferred given direct transmission between the two individuals. Next, we fit a statistical model to evaluate the extent to which epidemiological, sampling, genetic and phylogenetic factors influence the outcome of the inference. Third, we developed a statistical model that predicts the likely success of identifying transmission direction under real-life conditions when only routinely collected data are available. Using this model, we provide a framework to suggest how the accuracy of determining the transmission direction can be incorporated in population-level transmission studies when likely direct transmission pairs have been identified.

## Methods

### Ancestral state reconstruction

We used publicly available HIV-1 sequence data from 112 transmission pairs for which the direction of transmission is known following epidemiological investigation and where at least five sequences were available per individual (8). For each pair in our base case analysis, we inferred a Maximum Likelihood (ML) phylogenetic tree under a general time-reversible nucleotide substitution site model with the addition of heterogeneity of substitution rates among sites using a parametric Gamma model (GTR+G) with IQ-Tree (15). Trees were rooted using a subtype-specific sequence from the Los Alamos database which was removed from the tree in downstream analysis using the function drop.tip from the package ape (Analyses of Phylogenetics and Evolution) (17, 18). For each ML tree, we calculated the probability, *p*_i_, that ancestral state reconstruction correctly identifies the transmitting partner in each pair, *i*, while using the partner’s role in the transmission event as states. That is, after labelling the tips of each tree as sampled from either the transmitter or the recipient partner, we estimated the state probabilities at the root that maximized the likelihood of observing the state distribution at the tips using a joint estimation procedure (i.e., calculating the most likely state for each internal node in the tree while integrating over all the possible states along the other nodes in proportion to their probability) and assuming equal rates of transition between the two states. These analyses were conducted using the function ace (Ancestral Character Estimation) from the R package ape (16, 17).

We conducted sensitivity analyses to assess the role of phylogenetic reconstruction in assigning the transmission direction. First we assessed the role of the branch lengths estimated with a parametric Gamma model by calculating *p*_i_ from ML trees built under a four category non-parametric rate heterogeneity model (FreeRates) (18, 19) using IQ-Tree (15). We then evaluated whether Bayesian approaches improve the accuracy of the inferences. For this, we used hierarchical Bayesian inference with MrBayes 3.2.7 (20, 21) and simultaneously calculated a distribution of trees and the corresponding ancestral state posterior probabilities at the root, X_*i*_; we then defined the probability density *p*_*i*_ *= Pr*(X_*i*_ > 0.5). Finally, we inferred the ancestral states of each ML tree using the most parsimonious reconstruction which, instead of providing state probabilities, selects the state at the root that incurs the smallest number of state changes that are needed to observe the state distribution at the tips. For this, we used the Sankoff algorithm with the function ancestral.pars (Ancestral character reconstruction) from the R package phangorn (Phylogenetic Reconstruction and Analysis) (22–24).

### Phylogenetic inference of transmission direction

In our base case analysis, we classified the inferred direction of transmission as “consistent” with the known transmission direction if *p*_*i*_ >0.5, or “inconsistent” otherwise. In a sensitivity analysis, we accounted for a third “equivocal” outcome by classifying the inferred direction of transmission for each transmission pair, *i*, as either “consistent” if *p*_i_ ≥*t*, “inconsistent” if *p*_i_ ≤ 1-*t*, or “equivocal” otherwise. We used both a relaxed threshold of *t*=0.6 and a conservative threshold of *t*=0.95 for this ordinal three-class outcome. For the parsimony-based approach, we classified the inferred direction of transmission as either “consistent” if the state at the root was the transmitting partner, “inconsistent” if the state at the root was the recipient partner, or “both” if both partners were equally parsimonious at the root.

### Explaining the accuracy of phylogenetic inference of transmission direction

We evaluated in what circumstances ancestral state reconstruction succeeds in identifying the transmitting partner. For this, we built a suite of logistic regression models to predict the inferred direction of transmission as a function of information available from all transmission pairs. That is, for the base case binary outcome, the probability that the inferred direction of transmission is consistent with the known transmission direction, while for the three-class outcome, the probability that the inferred direction of transmission is consistent or inconsistent with the known transmission direction. We used 13 covariates as predictor variables that we organized into four classes: Epidemiological, Sampling, Genetic and Phylogenetic (**Table 1**).

**Table 1.** Covariates used in the two models

| Class | Covariate | Values (units where applicable) * |  |
| --- | --- | --- | --- |
|  |  | Model with all data | Model with routinely available data |
| Epidemiological information (E) | Sexual risk exposure group | Men-who-have-sex-with-men (MSM) or Heterosexual (HET) |  |
|  | Recency of the transmitter's infection | Acute (transmission occurred up to 90 days after the transmitter's infection), or *Chronic* (otherwise) | Excluded |
| Sampling information (S) | Sample size | Low (number of unique sampled sequences in at least one the partners is less than 10) or *High* (otherwise)<br>- |  |
|  | Sample size difference | Difference* in the number of unique sampled sequences between the partners | Absolute difference in the number of unique sampled sequences between the partners |
|  | Time from transmission | Sum of the absolute time-to-sampling of both partners relative to transmission time (days) | Excluded |
| Genetic information (G) | Sequence alignment length | Number of base pairs |  |
|  | Intra-host nucleotide diversity difference | Difference* between the within-partner mean pairwise sequence diversity (substitutions per site) | Absolute difference between the within-partner mean pairwise sequence diversity (substitutions per site) |
|  | Multiplicity of infection | *Single* (probability of one founder unique sequence in the recipient is greater than or equal to 0.75) or<br>*Multiple* (otherwise)<sup>†</sup> | Excluded |
| Phylogenetic<br>information<br>(P)** | Topology class | Paraphyletic-polyphyletic (PP), Paraphyletic-monophyletic (PM) or<br>Monophyletic-monophyletic (MM)<sup>‡</sup> |  |
|  | Phylogenetic diversity<br>difference | Difference* between the sum of the<br>branch lengths of each partner<br>subtree<sup>§</sup> (substitutions per site) | Absolute difference between the<br>sum of the branch lengths of each<br>partner subtree<sup>§</sup> (substitutions per<br>site) |
|  | Root-to-tip difference | Difference* between the minimum<br>root-to-tip distances of the<br>partners' sequences substitutions<br>per site) | Absolute difference between the<br>minimum root-to-tip distances of<br>the partners' sequences<br>(substitutions per site) |
|  | Most basal tip identity | *Transmitter, Recipient or Both*. The<br>identity of the tip(s) that minimizes<br>the number of internal nodes along<br>the paths between itself and the<br>root | *Agree, Disagree or Ambiguous*.<br>Whether the identity of the tip(s)<br>that minimizes the number of<br>internal nodes along the paths<br>between itself and the root matches<br>the identity of the individual with<br>the higher ancestral state<br>probability at the root |
|  | Inter-host patristic<br>distance | The shortest patristic distance<br>between tips from the partners<br>(substitutions per site) | The shortest patristic distance<br>between tips from the partners<br>(substitutions per site) |

<sup>†</sup> As in (8)
<sup>‡</sup> As in Romero-Severson et al, 2016
<sup>§</sup> We used the sum of the edge lengths that give rise to only one tip in the subtree as in (25)
\* Subtraction of the recipient's value from the transmitter's value.
\*\* Illustrated in Supplementary **Supplementary Figure 1**; to build these covariates when using a posterior distribution of trees, we selected either the most frequent observation (in the case of qualitative covariates) or the mean shift mode (in the case of the quantitative covariates).

We first fitted the model with all 13 covariates from across the four classes to identify the best set of inferred direction of transmission predictors. Then, as a sensitivity analysis, we fitted an additional 15 models built from all possible combinations of these four classes. That is, four models with three classes (ESG, ESP, SGP and EGP), six models with two classes (EG, ES, SG, SP, GP and EP) and four single-class models (E, S, G and P). Each class includes all of the available covariates within that class. This class-based sensitivity analysis allows us to explore the qualitative importance of different sources of information to the inference of transmission direction separately and to help validate our results.

### Increasing the accuracy of transmission direction inference

The previous suite of statistical models assumes knowledge of the transmitter and recipient’s identity in addition to epidemiological information not typically known. To evaluate how to interpret the inferred direction of transmission under ‘real-life’ conditions when this information is unknown, we developed a second suite of models with a reformulated set of eight covariates which are described in **Table 1**.

To evaluate how transmission pair characteristics influence the accuracy of inferring the transmission direction, we created datasets with observations that represent all possible combinations of the eight covariate values. For the discrete covariates we used the original categorical values, while for the continuous covariates we used a range of values evenly distributed between the minimum and the maximum of the original data. Using the best model from the binary suite of models and the best model from each of the ordinal suites of models (*t*=0.6 or *t*=0.95), we calculated the probability of inferred direction of transmission across the range of transmission pair characteristics.

### Model fitting, comparison and selection

We fitted all statistical models using least absolute shrinkage and selection operator (Lasso) regression with the function glmnet (fit a GLM with Lasso or elastic-net regularization) from the R package glmnet (Lasso and Elastic-Net Regularized Generalized Linear Models) (26) for the binary models and with the function ordinalNet (Ordinal regression models with elastic net penalty) from the R package ordinalNet (Penalized Ordinal Regression) (27) for the ordinal models. Using this approach we reduced overfitting because the resulting coefficients can be interpreted as evidence against the inclusion of a covariate if the coefficient shrinks to zero (28). The shrinkage coefficient was estimated using leave-one-out cross validation which provides an unbiased estimate of model performance when the dataset is small and, in particular, when one of the outcomes is uncommon and the data cannot be partitioned into training and validation sets. To compare the binary models, we calculated the area under the curve (AUC) statistic with the function auc (Compute the area under the ROC curve) from the R package pROC (Display and Analyze ROC Curves) (29). For ordinal models, we calculated a macro-AUC by averaging all results (one versus the rest) with linear interpolation between points using the the package function multi_roc (Multi-class classification ROC) from the R package multiROC (Calculating and Visualizing ROC and PR Curves Across Multi-Class Classifications) (30). We considered models with AUC > 0.9 to have high discriminatory power and selected the best ranking models as those with the highest AUC within three decimal places.

## Results

### Data

The 112 transmission pairs exhibited wide variation across all the epidemiologic, sampling, genetic, and phylogenetic characteristics evaluated (**Supplementary Figure 2**). Specifically, most of the transmitters were in the chronic stage at the time of transmission (101/112) pairs and most of the transmitters were reported as HET (36/112). The sample size was low (i.e., fewer than 10 sequences in the least sampled individual within the pair) in 60/112 pairs, while the median sample size difference was 5.5 unique sequences (interquartile range—IQR—1.00-13.25), and the median sampling time of both partners relative to the recipient time of infection was 173 days (IQR 84-410 days).

The median sequence alignment length was 1,534 base pairs (IQR 747-2,591); a total of 103/112 of the pairs had sequences that spanned the *env* region while 9/112 spanned the *gag* region; the median difference of intra-host nucleotide diversity was 0.013 substitutions per site (IQR 0.005-0.030), while 84/112 recipient’s infections were more probably seeded by a single variant.

The most frequent topology class was PM (62/112), followed by PP (30/112) and MM (20/112), while the median difference in phylogenetic diversity was 0.051 substitutions per site (IQR 0.010-0.122), the median difference in minimum root-to-tip distances was 0.007 substitutions per site (IQR 0.002-0.020) and the median of the minimum inter-host patristic distance was 0.009 substitutions per site (IQR 0.003-0.020). In terms of the most basal tip identity, the tip closest to the root (i.e., the one separated by the least number of internal nodes) belonged to the transmitter partner in 86/112 pairs, to the recipient partner in 12/112 pairs, and tips from both partners were equally closer to the root in 14/112 pairs.

### Phylogenetically inferred direction of transmission

We found that probabilistic ancestral state reconstruction on phylogenetic trees tends to correctly infer the transmission direction, with 83.9% (94/112) of the pairs being consistent (*p*_*i*_ >0.5) and 16.1% (18/112) of the pairs inconsistent with the known transmission direction (**Figure 1, Supplementary Table 1**). There were significant differences in the topology class by outcome (Pearson’s Chi-squared *P* < 0.001). When the transmission direction was correctly inferred, the PM topology class predominated (59/94); in contrast, when the transmission direction was incorrectly inferred, MM and PP topology classes predominated (8/18 and 7/18, respectively).

**Figure 1.**
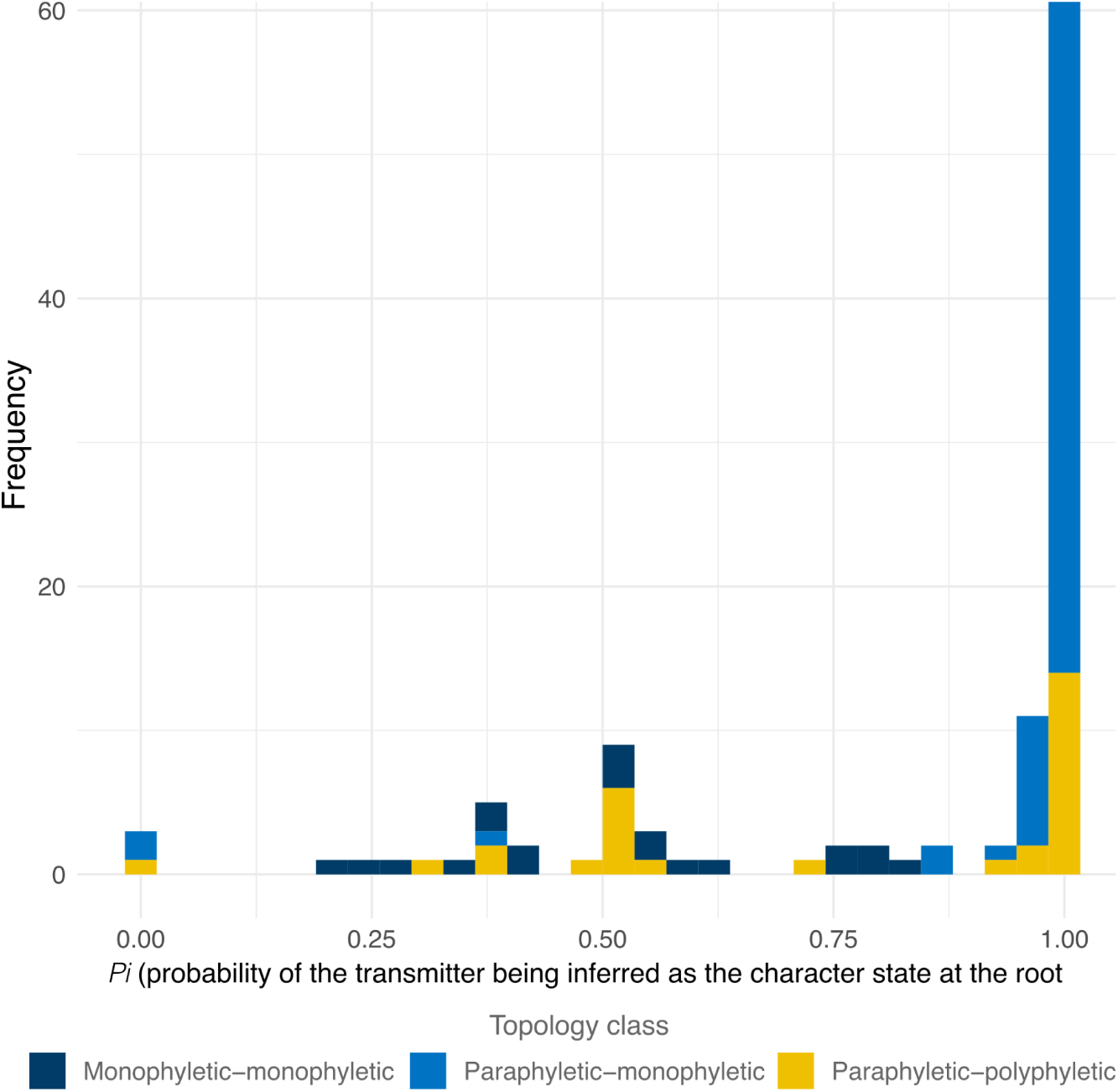
The probability for each transmission pair, *i*, that the transmitting partner is correctly identified using ML ancestral state reconstruction. Observations are colored by the topology class. Observations with *p*_*i*_ >0.5 indicated that the inferred transmission direction was consistent with the known transmission history.

### Explaining the accuracy of phylogenetic inference of transmission direction

The AUC characterises the probability of discriminating between the correct and incorrect transmission direction. We found that the 16 logistic models varied greatly in their discriminatory power to detect when the phylogenetically-inferred transmission direction was correct given that the AUC values ranged between 0.723 and 0.976 (**Figure 2A**). There were seven models with an AUC greater than 0.9, with a median AUC of 0.974 and with little separating their discriminatory power (the maximum ΔAUC was 0.006); these seven models all included at least four out of five covariates from the phylogenetic class (P) after variable selection and regularization (**Figure 2B**).

**Figure 2.**
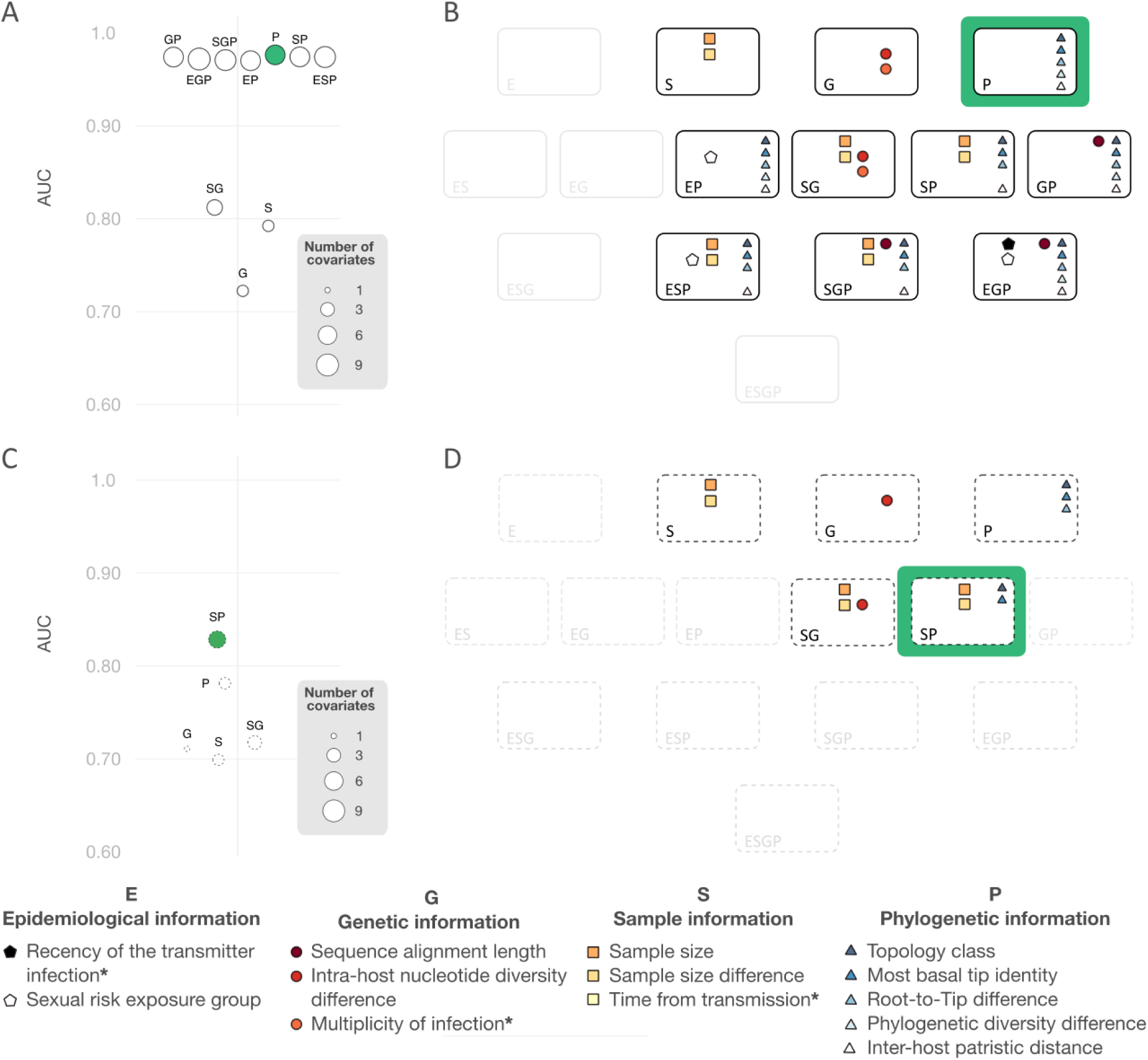
Model results. (A) AUC of the models (represented by circles). The name of the model indicates the class of information included in the model (i.e. Epidemiological, Genetic, Sample or Phylogenetic). The size of each circle indicates the number of covariates that were kept in the model after Lasso regression; the model with the highest AUC is highlighted in green. (B) The subset of covariates that were included in each model after Lasso regression, colored by class. The number of covariates inside models in panel B corresponds to the size of the models in panel A. Grayed-out models indicate those for which Lasso regression returned either a null model or a model without covariates from all the included classes. (C) and (D) as in the top panels but using only covariates that are routinely available and where the known direction of transmission was not considered during the definition of these covariates. The three covariates that were excluded in this second analysis are marked with ‘*’.

The model P kept all the five covariates and had the highest discriminatory power (0.976, **Supplementary Table 2**). In this model, the probability of correctly inferring the transmission direction increases (i) when we observe a PM or a PP topology class (compared to MM), (ii) when the most basal tip in the tree corresponds to a sample from the transmitter (compared to a tree when tips from both partners are equally basal) (iii) when the phylogenetic diversity of the transmitter is larger than that of the recipient, (iv) the root-to-tip distance of the recipient is larger than than of the transmitter, and (v) when the minimum inter-host patristic distance gets larger. In contrast, the probability of correctly inferring the transmission decreases when the most basal tip corresponds to a sample from the recipient (compared to a tree when tips from both partners are equally basal).

### Increasing the accuracy of transmission direction inference

#### Base case analysis

While the models inform about the covariates that affect the phylogenetic inference of the transmission direction between a pair of individuals where direct transmission has been previously established they do not inform about the accuracy of the inference when we don’t know who infected whom. To tackle this issue, we re-analyzed the data after masking the identity of the transmitter and recipient and using only routinely available information. In our base case analysis (a binary model when the outcome is consistent if *p*_*i*_ >0.5, inconsistent otherwise), the model fitting reduced the number of models to a total of five that were either single-class models (S, G, P) or the dual-class models SG and SP (**Figure 2C**). Model SP was the best-fitting model (AUC = 0.827), with sample size, sample size difference, topology class, and the identity of the most basal tip, being the covariates (**Figure 2D**). Model SP suggests that the probability that the transmitting partner is correctly identified is higher when one of the virus populations is small and embedded as a monophyletic group in a much larger virus population of the partner. Specifically, (i) when the sample size is large (compared to small), (ii) when the difference in sample size gets larger, (iii) when we observe a PM topology class (compared to either MM or PP), and (iv) when the identity of the most basal tip agrees (compared to either disagreeing or being ambiguous) with the identity of the individual with the highest probability at the root.

#### Sensitivity analyses

##### Equivocal outcomes

When we classified the inferred direction of transmission to be either consistent, inconsistent or equivocal with a relaxed probability threshold (*p*_*i*_ ranging between 0.4 and 0.6 as equivocal), the inferred direction of transmission was consistent with the known transmission direction in 74.1% (n=83) of the pairs, equivocal in 13.4% (n=15) and inconsistent in 12.5% (n=14) (**Supplementary Table 1**). Similarly, using a conservative threshold (*p*_*i*_ ranging between 0.05 and 0.95 as equivocal) increased the proportion of pairs that are classified as equivocal to 33.0% (n=37) and reduced the proportions of consistent and inconsistent pairs to 64.3% (n=72) and 2.7% (n=3), respectively. Regardless of the threshold, the ordinal model with the highest macro-AUC was model P (the phylogenetic class only model), but the discriminatory power of this model was lower for the conservative threshold than that of the relaxed one (AUC = 0.765 vs. 0.843 respectively (**Supplementary Figure 3, Supplementary Table 3**). Similar to the base case binary model, the ordinal models show that the probability of the inferred direction of transmission being correct is higher when one of the virus population is embedded as a monophyletic group in the virus population of the partner. In the model, this refers to when we observe a PM topology (compared to either MM or PP), and when the identity of the basal tip agrees (compared to either disagree or ambiguous) with the identity of the individual with the highest probability at the root. In addition, this probability increases when the phylogenetic diversity in one of the individuals is greater to that of the partner and, in the case of the conservative threshold, also increases when only one of the partners is close to the root and when viral populations have not diverged much (the minimum inter-host patristic distance gets smaller).

#### Most parsimonious ancestral state reconstruction

When we used the most parsimonious reconstruction to calculate the inferred transmission direction from the ML trees, the model SP had the highest macro-AUC and an equivalent discriminatory power than the best-case scenario of the probabilistic ordinal approach (AUC = 0.844; ΔAUC of 0.004) (**Supplementary Figure 3, Supplementary Table 3)**. This model suggests that the probability of the inferred direction of transmission being correct increases as the difference in sample size gets larger, when we observe a PM topology (compared to either MM or PP), and when the identity of the most basal tip agrees (compared to either disagreeing or being ambiguous) with the most parsimonious state at the root.

#### Tree reconstruction methods

When we used either ML tree reconstruction with a non-parametric rate heterogeneity model (R4) or Bayesian inference under a GTR+G4 model, we found that the binary model with the highest macro-AUC was the model GP (AUC of 0.853 and 0.867 for ML+R4 and Bayesian inference, respectively). On the other hand, with ML under GTR+R4 model the best ranking ordinal model was model P regardless of the threshold (an AUC of 0.835 and 0.821 for *t*=0.6 and *t*=0.95, respectively). For Bayesian inference, the top ranked models were model P with t=0.6 and model SP with t=0.95 with an AUC of 0.837 and 0.747, respectively (**Supplementary Figure 4, Supplementary Table 3**). The GP and SP models included the covariate difference in intra-host nucleotide diversity and difference in the number of sampled unique sequences, respectively. All GP and P models included the two topological covariates, that is the topology class and whether the identity of the most basal tip agrees or disagrees with the identity of the individual with the higher probability at the root. When the equivocal outcome was considered, the covariates that rely on branch lengths (i.e., phylogenetic diversity, root-to-tip and patristic distances) became additional predictors for the correct identification of the transmission direction with the same effects as for the base case ordinal models, with the exception of the model built under ML with GTR+R4 and t=0.6 in which only the two topological covariates remain as important.

### Implications for bias within population studies

We next evaluated whether routinely undisclosed epidemiological characteristics are associated with the probability of correctly identifying the direction of transmission. Specifically, we found that the stage of the transmitter’s infection at the time of transmission is associated with the topology class of the phylogenetic tree. That is, PP topologies—that are associated with less chance of accurately predicting the transmitting partner—are more frequently observed (72.7%) when transmission occurred during the transmitter’s acute stage and PM topologies—that are associated with more chance of accurately predicting the transmitting partner—are more frequently observed (58.4%) when transmission occurred during the transmitter’s chronic stage (Pearson’s chi-squared test *P* < 0.001). Therefore, because the stage of the transmitting partner’s infection is likely to influence the topology class of the phylogenetic tree, which, in turn influences the probability of correctly identifying the transmitting partner, there is a risk of overrepresentation chronic stage infections in the set of correctly identified transmission pairs.

### Implications for inference of transmission direction

Our analysis suggests that transmission pair characteristics influence the likelihood of correctly identifying the transmission direction using ancestral state reconstruction. To estimate the practical importance of this result, we used the best binary and ordinal models (SP and P) to predict the chance of inferring the correct transmission direction across all possible covariates.

Our results suggest that, in our binary and ordinal model with an equivocal threshold of 0.6, observing a PM topology is sufficient to provide at least a 75% chance of correctly identifying the transmitting partner (**Figure 3**). If the identity of the most basal tip agrees with the identity of the individual with the higher state probability at the root, this chance increases to at least 90%. If the classification is between consistent or inconsistent, this probability further increases by observing a minimum difference in the number of unique sequences in the samples. (**Figure 3A**). Conversely when the classification is between consistent, inconsistent and equivocal, this probability further increases by observing a minimum difference in phylogenetic diversity.

**Figure 3.**
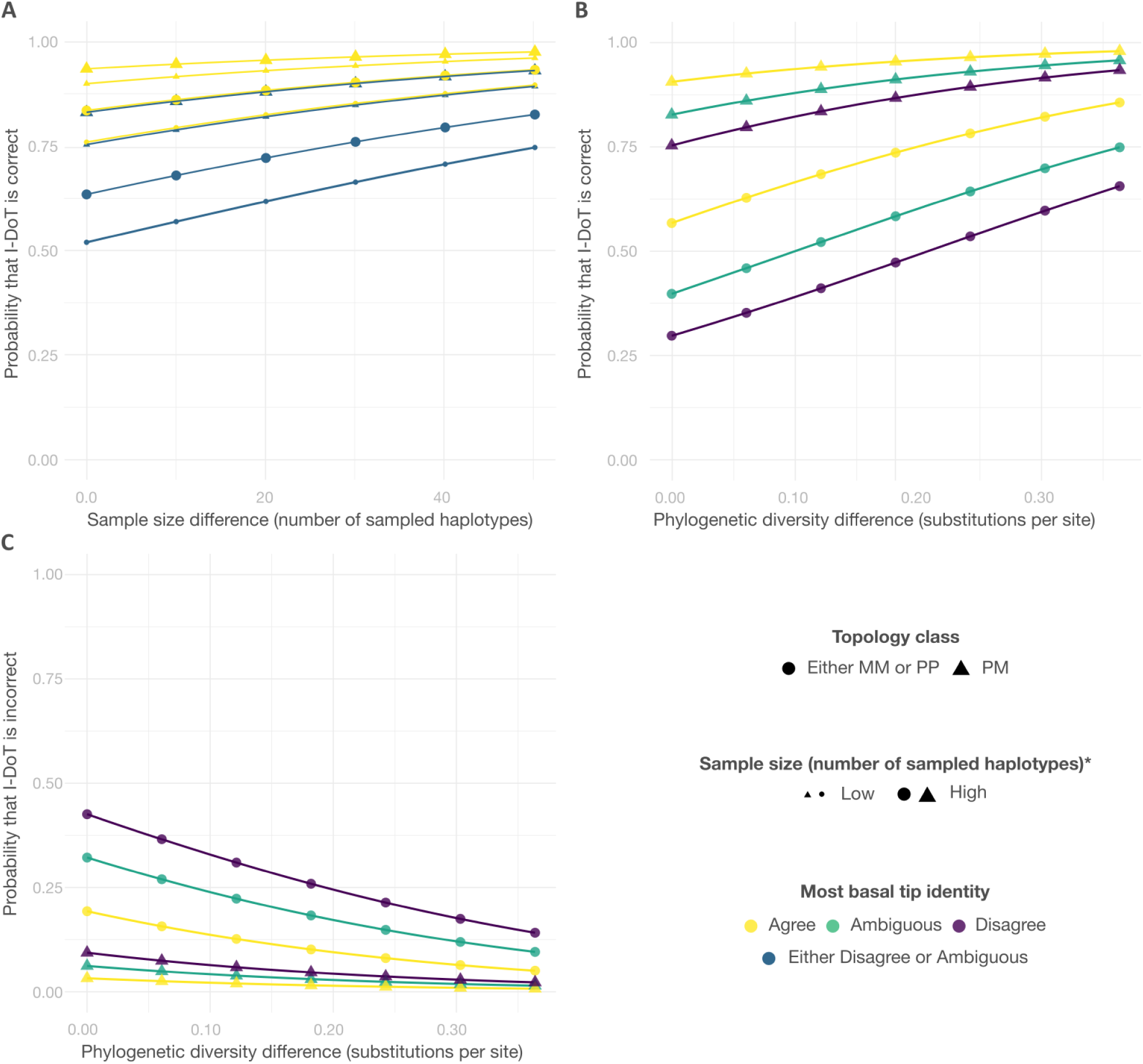
Predicting the success of inferring the direction of transmission. (A) The binary ‘SP’ model with four predictor covariates. (B) and (C) ordinal model ‘P’ model (with relaxed threshold for direction of transmission classification *t*=0.6) with three predictor covariates. (*) Sample size only applies to binary ‘SP model’ in (A).

## Discussion

We have combined empirical data on well-characterized HIV transmission pairs with statistical modelling to determine the conditions under which probabilistic phylogenetic analysis correctly infer the direction of HIV transmission. Our results suggest that, while ancestral state reconstruction correctly identifies the transmission direction in the majority of known transmission pairs, this success is determined by the epidemiological, sampling, genetic and phylogenetic characteristics of the individuals and their viral populations. We show that topological and branch-length metrics—such as root-to-tip distances—from the phylogenetic tree of the transmission pair, affect the chances of successfully inferring the transmission direction.

To guide future work on identifying the transmitting partner within a linked HIV pair, we quantified the probability of correctly inferring the transmission direction as a function of readily obtainable information. Under these circumstances, a PM topology and a match between the identity of the tip closer to the root (i.e., the one separated by the least number of internal nodes) and the identity of the state assigned to the root were highly predictive of inferring the correct transmission direction. This result agrees with the theoretical prediction that when multiple viral sequences per individual are available, the relative ordering of sequence clusters from the two individuals should inform transmission direction inference (9). Moreover, our results suggest that using a relative metric of the difference in intra-host diversity between the partners improved discriminatory power (with larger differences indicative of a greater chance of correctly identifying the transmission direction), which is consistent with previous work (12, 13).

There is a noticeable drop in discriminatory power when our models only include readily obtainable data, which is likely due to the loss of discriminatory information. Indeed, two variables that are not typically known and are not included under our real-life conditions model—the recency of the transmitter’s infection at the time of transmission and the time from transmission—have been shown to influence the topology class (8, 9), which in turn influences the chance of correctly identifying the transmitting partner. In the absence of such data, our results confirm that inferences about directionality can entail considerable uncertainty (31). Nonetheless, our results shed light into the possible reasons for variability between studies. For instance, studies that most successfully infer the transmission direction used next generation sequencing data from heterosexual serodiscordant couples, where transmission occurs during the chronic stage of the transmitter (12, 14) and there is a higher likelihood of a PM topology that our model suggests is indicative of correctly identifying the transmission direction.

Here we show that even when we are conservative about attributing the transmission direction using ancestral state reconstruction, requiring 95% of the trees in a distribution to support a direction, a small percentage of cases still have an incorrect prediction. A recent study tested whether the prediction of the transmission direction could be improved by using NGS and in the best-case scenario the prediction was incorrect for 4/33 (12%) pairs (13). Nonetheless, easily calculated measures of relative intra-host diversity can increase the probability that the direction of transmission is correctly identified (the difference in the numbers of unique sequences, the difference in mean pairwise nucleotide diversity, the difference in phylogenetic diversity or the minimum root-to-tip distance). Our results suggest that these metrics can be evaluated for each pair concurrently with ancestral state reconstruction, which would provide a probability of incorrectly inferring transmission direction. In turn, these probabilities could be used to either select a subset of pairs for which a smaller probability of incorrect assignment is likely, or as weights to adjust further analysis which could be useful to move beyond identifying potential transmission pairs or clusters and allow to interpret transmission inferences by considering the degree of uncertainty in the transmission direction.

Our results suggest that there was little difference in the ordinal classification performance of ancestral state reconstruction methods (either probabilistic or parsimony-based algorithms) when a Maximum Likelihood tree is used. However, we did find differences in the nature of the data that were able to predict whether the inference was correct. That is, while differences in the number of sampled unique sequences remain high using a parsimony-based approach, only phylogenetic information is required for probability-based approaches. These differences likely occur because probabilistic methods, unlike parsimony-based algorithms, incorporate information about branch lengths during the inference, which are indicators of nucleotide diversity.

This study has some limitations: first well characterized transmission pairs are scarce, and we were not able to test our models out of our sample so we relied on leave-one-out cross validation to minimize overfitting and avoid biased estimates of model performance. Second, we used relational metrics that summarized the magnitude of differences in the intra-host diversity (i.e., differences in the number of sampled unique sequences, in intra-host nucleotide diversity, and in minimum root-to-tip distance), in the inter-host-diversity (i.e., differences in minimum patristic distance), or in a composite measure of diversity (e.g. differences in phylogenetic diversity). However, there are alternative ways to conceptualize diversity and there may be other factors that affect the correct inference of transmission direction while not represented in the available data. Third, we did not consider the effects of processes such as superinfection and recombination, which impact on diversity and phylogenetic interpretation. Finally, we considered only pairs of individuals for whom direct transmission had been verified using information such as contact tracing and testing histories as detailed elsewhere (8). While these data are currently the best available, it is conceivable that there may have been an unsampled intermediate partner or common source partner.

Our study only addresses the chance of correctly identifying the transmission direction given that the direct transmission of infection between the two individuals has been previously established. In reality, we would also like to identify direct infection transmissions between individuals, and to rule out transmission through one or more intermediate partners or common sources. However, this latter question is inherently more complex because not only is the range of possible outcomes larger, but also because the data on which to fit a model is scarce. Moreover, while arguably phylogenetic methods serve as an important tool to understand infectious disease epidemiology, they are unsuitable for individual pair-level analysis such as those involved in forensics because these methods entail considerable uncertainty.

While the use of phylogenetic analysis to infer the direction of transmission has recently shown promise, there has been considerable variation in accuracy across studies. Here we provide a statistical framework to help explain these differences and to improve the reliability in future work. We stress that while phylogenies provide rich and important information about transmission, conclusions on directionality must be considered cautiously and with full adherence to the strictest ethical standards of data use.

## Data Availability

This study uses publicly available genetic and epidemiological data that was generated in previous studies and that was collated and described in 10.1126/science.aba5443. The data can be retrieved from The Los Alamos HIV and GenBank databases

## Acknowledgements

We are grateful to Art Poon for helpful feedback. CJVA and KEA were funded by an ERC Starting Grant awarded to KEA (award number 757688). MH was funded by The HIV Prevention Trials Network (grant number H5R00701.CR00.01) and The Bill and Melinda Gates Foundation (grant number OPP1175094). JACB was supported by the MRC Precision Medicine Doctoral Training Programme (ref: 2259239). KAL was supported by The Wellcome Trust and The Royal Society grant no. 107652/Z/15/Z. JB received support from the UK MRC and the UK DFID (#MR/R010161/1) under the MRC/DFID Concordat agreement and as part of the EDCTP2 Programme supported by the European Union.

