## Supplementals for "USING PHYLOGENETICS TO INFER HIV-1 TRANSMISSION DIRECTION BETWEEN KNOWN TRANSMISSION PAIRS"

**Supplementary Table 1. Inferred Direction of Transmission (I-DoT) by method**

| I-DoT | Maximum Likelihood | | | | | | | Bayesian Inference | | |
|---|---|---|---|---|---|---|---|---|---|---|
| | GTR+G | | | | GTR+R | | | GTR+G | | |
| | Binary | Multi-categorical | | | Binary | Multi-categorical | | Binary | Multi-categorical | |
| | t=0.5 | t=0.60 | t=0.95 | MPR† | t=0.5 | t=0.60 | t=0.95 | t=0.5 | t=0.60 | t=0.95 |
| Consistent | 94 (83.9%) | 83 (74.1%) | 72 (64.3%) | 80 (71.4%) | 92 (82.1%) | 84 (75.0%) | 70 (62.5%) | 98 (87.5%) | 89 (79.5%) | 69 (61.6%) |
| Equivocal | NA | 15 (13.4%) | 37 (33.0%) | 26 (23.2%) | NA | 16 (14.3%) | 38 (33.9%) | NA | 16 (14.3%) | 39 (34.8%) |
| Inconsistent | 18 (16.1%) | 14 (12.5%) | 3 (2.7%) | 6 (5.4%) | 20 (17.9%) | 12 (10.7%) | 4 (3.6%) | 14 (12.5%) | 7 (6.2%) | 4 (3.6%) |

† Most parsimonious reconstruction
I-DoT: inferred direction of transmission

**Supplementary Table 2. Details of the base-case top-ranked classification model with all data**

| **Tree-Inference method*** | **Site-model** | **Strategy** | **Threshold** | **Model** | **AUC** | **Covariates** [level][†] (Shrinkage coefficient**)** |
|---|---|---|---|---|---|---|
| Maximum Likelihood | GTR+G | Binary | t=0.5 | P | 0.976 | Topology class[PM] (0.708)<br>Topology class [PP] (0.025)<br>Root-to-tip difference (-467.576)<br>Phylogenetic diversity difference (7.343)<br>Most basal tip identity [source] (1.091)<br>Most basal tip identity [recipient] (-0.380)<br>Inter-host patristic distance (26.540) |

[†] Level for discrete covariates

### Supplementary Table 3. Details of the top-ranked classification models with routinely-available data

| Tree-Inference method | Site-model | Strategy | Threshold | Model | (Macro) AUC | **Covariates** [level][†] (Shrinkage coefficient) |
|---|---|---|---|---|---|---|
| Maximum Likelihood | GTR+G | Binary | t=0.5 | SP | 0.826 | Sample size [low] (-0.472)<br>Sample size difference (0.020)<br>Topology class [PM] (1.0371)<br>Most basal tip identity [agree] (1.068) |
| | | Multi-categorical | t=0.60 | P | 0.843 | Topology class [PM] (1.968)<br>Phylogenetic diversity difference (4.137)<br>Most basal tip identity [agree] (0.684)<br>Most basal tip identity [disagree] (-0.445) |
| | | | t=0.95 | P | 0.765 | Topology class [PM] (2.268)<br>Root-to-tip difference (10.263)<br>Phylogenetic diversity difference (3.849)<br>Most basal tip identity [agree] (1.138)<br>Inter-host patristic distance (-16.152) |
| | | | Most parsimonious reconstruction | SP | 0.844 | Sample size difference (0.042)<br>Topology class [PM] (1.288)<br>Most basal tip identity [agree] (2.370) |
| | GTR+R | Binary | t=0.5 | GP | 0.853 | Intra-host nucleotide diversity difference (11.107)<br>Topology class [PM] (1.560)<br>Phylogenetic diversity difference (0.270)<br>Most basal tip identity [agree] (0.788) |
| | | Multi-categorical | t=0.60 | P | 0.835 | Topology class [PM] (1.639)<br>Most basal tip identity [agree] (0.422) |
| | | | t=0.95 | P | 0.821 | Topology class [PM] (2.026)<br>Phylogenetic diversity difference (1.447)<br>Most basal tip identity [agree] (1.115) |
| Bayesian Inference | GTR+G | Binary | t=0.5 | GP | 0.867 | Intra-host nucleotide diversity difference (37.817)<br>Topology class [PM] (0.851)<br>Most basal tip identity [agree] (0.329)<br>Most basal tip identity [disagree] (-1.749) |
| | | Multi-categorical | t=0.60 | P | 0.837 | Topology class [PM] (1.699)<br>Phylogenetic diversity difference (5.480)<br>Most basal tip identity [agree] (1.160) |
| | | | t=0.95 | SP | 0.837 | Topology class [PM] (1.563)<br>Root-to-tip difference (23.247)<br>Most basal tip identity [agree] (0.789)<br>Inter-host patristic distance (-10.225) |

[†] Level for discrete covariates

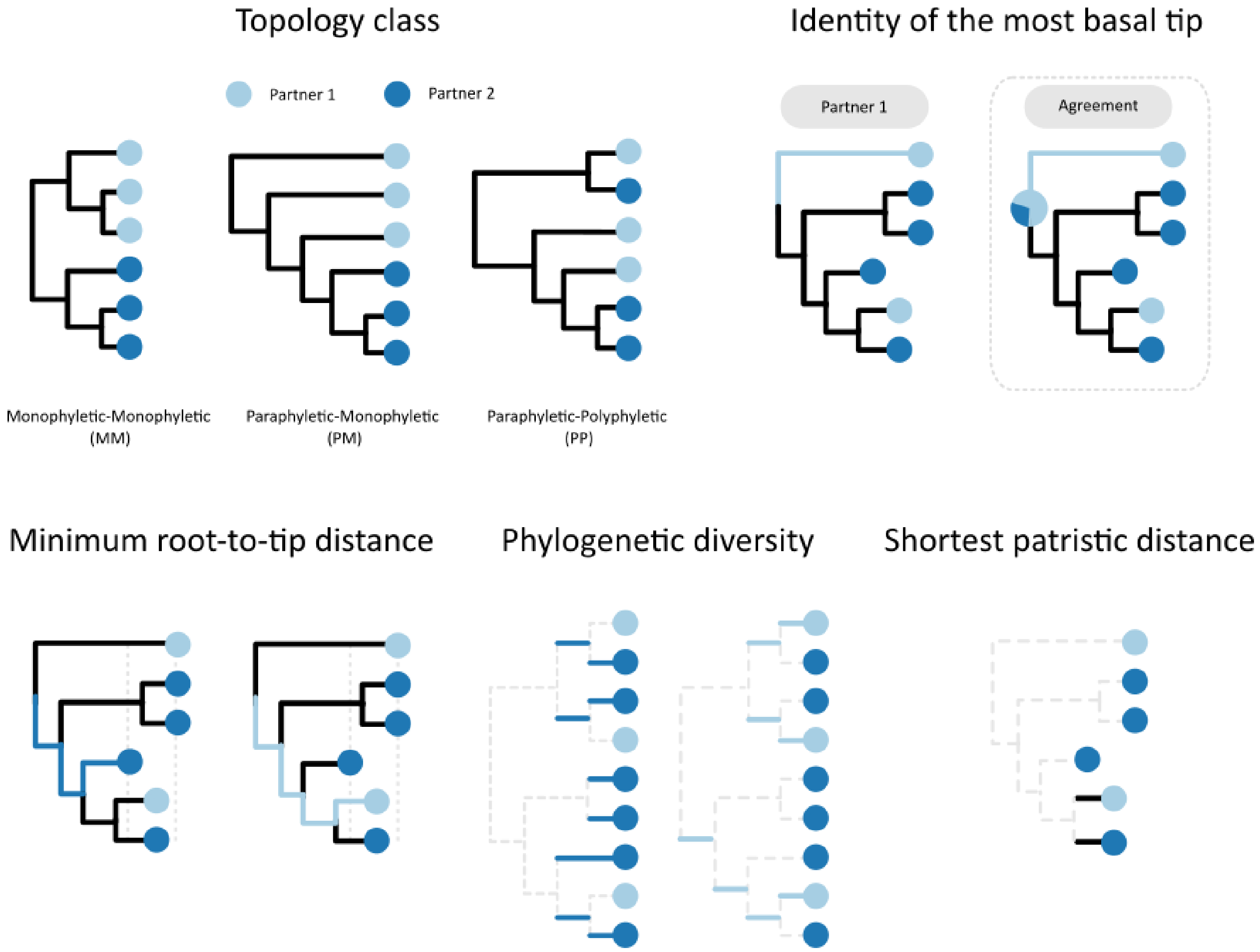


**Supplementary Figure 1.** Illustration of the different metrics that are used to define the covariates from the *phylogenetic information* class. The *topology class*, either paraphyletic-polyphyletic (PP), paraphyletic-monophyletic (PM) or monophyletic-monophyletic (MM). The *identity of the most basal tip*, *i.e.* the identity of the tip that minimises the number of internal nodes along the paths between the root and the tips (the alternative definition—inside the square—corresponds to the identity of the most basal tip when it agrees or disagrees with the identity of the individual with the higher probability at the root). The *minimum root-to-tip distance*, i.e. the shortest path from the root to the tips of an individual (calculated for each partner). The *phylogenetic diversity* using the unique evolutionary history measure, i.e. the sum of the branch lengths that are not shared across the subtree of an individual and which give rise to each single tip of the individual (calculated for each partner), as described in the function pd.calc from the R package Caper. The *shortest patristic distance* between the tips of the two partners, i.e. the shortest path connecting a tip from both individuals.

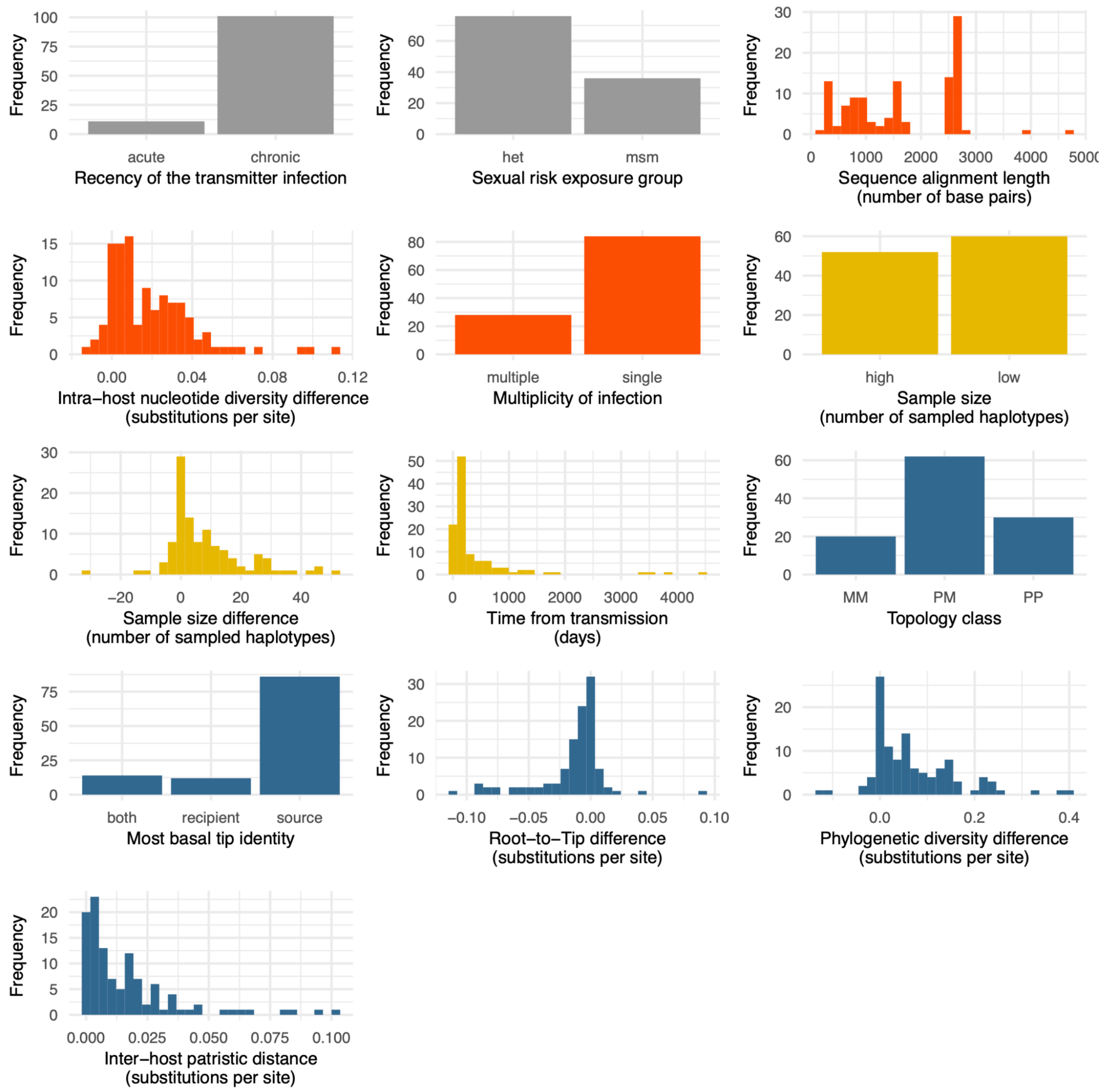


**Supplementary Figure 2.** Distribution of the covariates values colored by covariate class: epidemiological (gray), sampling (coral), genetic (dull yellow) and phylogenetic (blue)

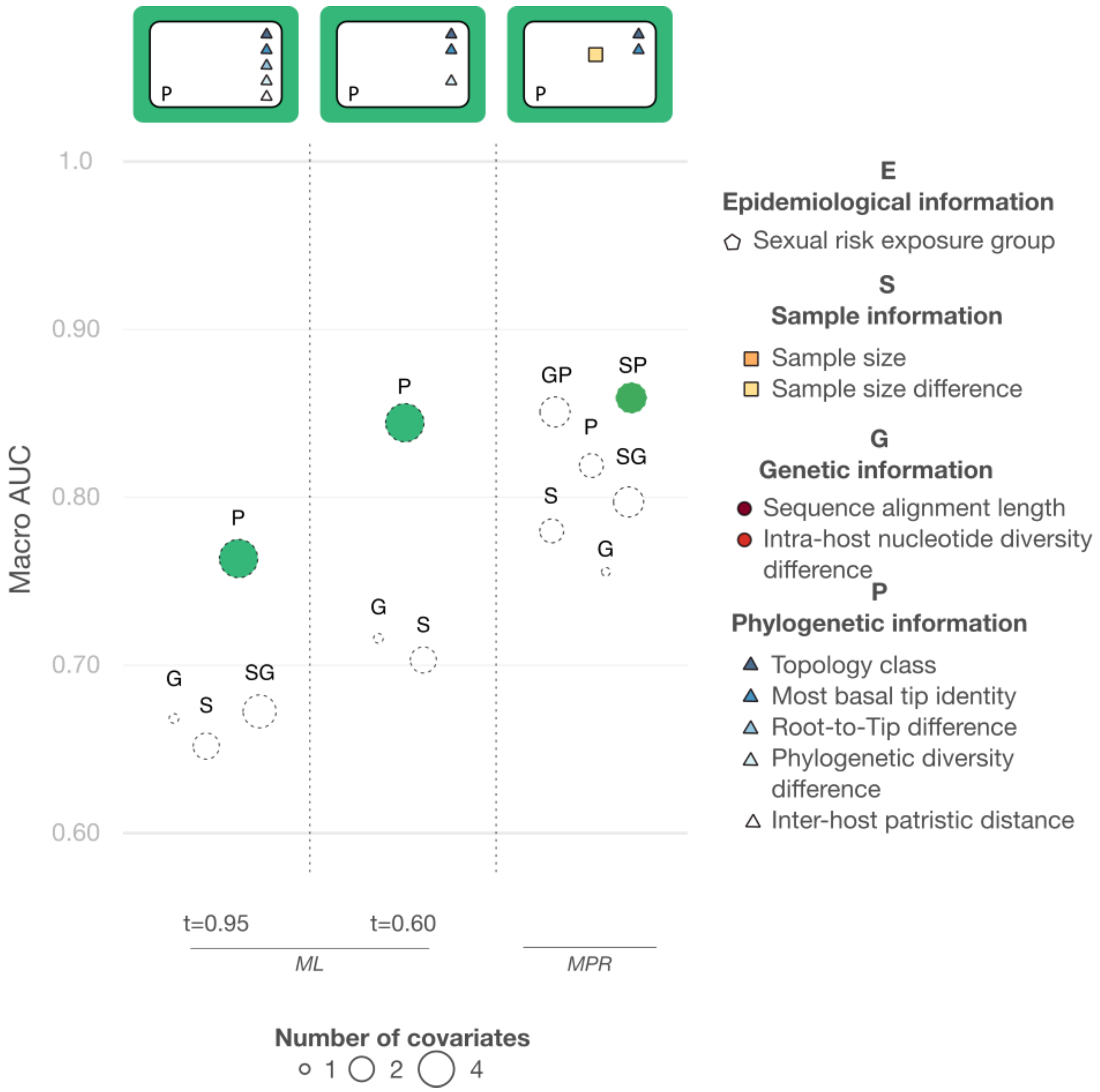


**Supplementary Figure 3. Ordinal models outcomes when using routinely-available data.** Macro-AUC of the multi-categorical models (represented by circles) using Maximum Likelihood (ML) or the Most Parsimonious reconstruction (MPR). The ML results are presented for the relaxed (*t*=0.60) and the conservative thresholds (*t*=0.95). The name of the model indicates the class of information included in the model (i.e. Epidemiological, Genetic, Sample or Phylogenetic). The size of each circle indicates the number of covariates that were kept in the model after Lasso regression. The green color fill underscores the models with the highest AUC. The top rectangles indicate the subset of covariates that were included in the models with the highest AUC after Lasso regression, colored by class; the number of covariates inside these rectangles corresponds to the size of the circles.

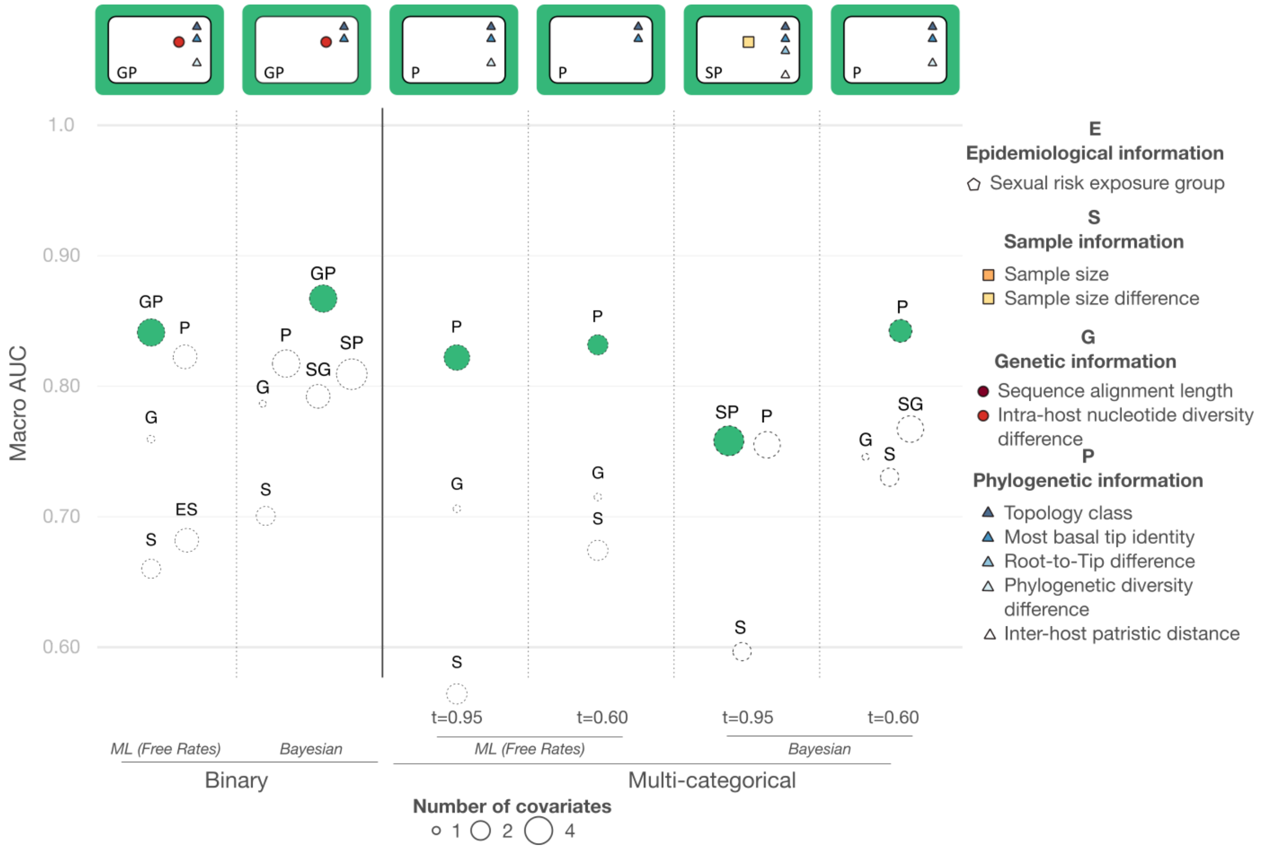


**Supplementary Figure 4. Binary and ordinal models outcomes when using routinely available data.** Macro-AUC of the models (represented by circles) using a Maximum Likelihood (ML) with FreeRates or using Bayesian Inference. In the multi-categorical scenarios, results are presented for the relaxed (*t*=0.60) and the conservative thresholds (*t*=0.95). The name of the model indicates the class of information included in the model (i.e. Epidemiological, Genetic, Sample or Phylogenetic). The size of each circle indicates the number of covariates that were kept in the model after Lasso regression. The green color fill underscores the models with the highest AUC. The top rectangles indicate the subset of covariates that were included in the models with the highest AUC after Lasso regression, colored by class; the number of covariates inside these rectangles corresponds to the size of the circles.
